# Considering the *APOE* locus in polygenic scores for Alzheimer’s disease

**DOI:** 10.1101/2019.12.10.19014365

**Authors:** Erin B. Ware, Jessica D. Faul, Colter M. Mitchell, Kelly M. Bakulski

## Abstract

Polygenic scores are a strategy to aggregate the small, additive effects of single nucleotide polymorphisms across the genome. With phenotypes like Alzheimer’s disease, which have a strong and well established genomic locus (*APOE)*, the cumulative effect of genetic variants outside of this area has not been well established in a population-representative sample. Here we examine the association between polygenic scores both with and without the *APOE* region at different *P* value thresholds. We also investigate the addition of *APOE*-ε4 carrier status and its effect on the polygenic score – dementia association. We found that including the *APOE* region through weighted variants in a polygenic score was insufficient to capture the large amount of risk attributed to this region. We recommend removing this region from polygenic score calculation and treating the *APOE* locus as an independent covariate.

## Introduction

The most common form of dementia is Alzheimer’s disease (AD), representing roughly 65% of dementia cases (Van Cauwenberghe, Van Broeckhoven, & Sleegers, 2016). AD is thought to arise from a combination of both genetics, environment, and lifestyle factors (Gatz et al., 2006). The estimated heritability of late onset Alzheimer’s disease is around 74% (Gatz et al., 1997). While large-scale genome-wide association studies (GWAS) have identified several genetic loci associated with AD (Harold et al., 2009; Jansen et al., 2019; Kunkle et al., 2019; Lambert et al., 2013; Naj et al., 2011; Seshadri et al., 2010; Tosto et al., 2017), being a carrier of the Apolipoprotein E (*APOE-ε4*) allele remains the strongest genetic predictor of AD (Ward et al., 2012). One copy of *APOE-ε4* (inheriting a CC at these two locations from either parent) confers a 3-fold risk of AD while two copies (inheriting a CC at these two locations from *both* parent) a 15-fold increase in risk (Liu, Liu, Kanekiyo, Xu, & Bu, 2013). The effect of *APOE-ε4* is all the more difficult to capture in a single variant GWAS as *APOE-ε4* is a haplotype composed of two SNPs: rs7412 and rs429358 – which will never be fully be captured in a traditional linear model GWAS framework. However, in GWAS many independent SNPs have been identified in and near the *APOE* gene locus. The *APOE* gene region contains many variants in high linkage disequilibrium within roughly 100 kilobases, including several additional high-risk sites in the translocase of outer mitochondrial membrane 40 (*TOMM40*) gene.

The largest AD GWAS meta-analysis to date (N=94,437) is the from the International Genomics of Alzheimer’s Project (IGAP) (Kunkle et al., 2019). This meta-analysis confirmed 20 previously identified AD risk loci (Lambert et al., 2013) and identified five new genome-wide loci including (*IQCK, ACE, ADAM10, ADAMTS1*, and *WWOX*). The IGAP used a three-stage strategy where Stage 1 consisted of genotyped and imputed data on 9,456,058 common and 2,024,574 rare single nucleotide polymorphisms (SNPs) to meta-analyze GWAS from four cohorts (n_cases_=21,982; n_controls_=41,944). Stage 2 included replication with a custom I-select genotyping chip developed in Lambert et al. 2013 (Lambert et al., 2013) and included 11,632 variants and 18,845 individuals with a meta-analysis of Stage 1 and Stage 2. Finally, Stage 3 replicated 44 variants and meta-analyzed Stages 1 and 2 and 3 for a total of 35,274 cases and 59,163 controls. The associations between millions of genetic loci and AD are documented in IGAP and available for testing in independent populations.

Many complex diseases may result from the consideration of small individual effects across the genome. Polygenic scores (PGS) are generally derived from the sum of weighted variants across an individual (International Schizophrenia et al., 2009; Martin, Daly, Robinson, Hyman, & Neale, 2019). Though conceptually simple, many analytic decisions contribute to different qualities of PGSs including different coefficient of variation (R^2^), correlations between scores, and areas under the curve. An important consideration for diseases such as Alzheimer’s disease – which have a genetic locus like *APOE/TOMM40* conferring much of the genetic risk to the disease – is to determine how the remaining variants in the genome contribute to the disease. Removing a region with many risk variants and deciding which and how many variants to include in a PGS can offer substantively different conclusions. For instance, one study reported a PGS area under the curve of 0.57 for Alzheimer’s disease (parental proxy) using 21 SNPs and excluding the *APOE* region (Tosto et al., 2017), while another study reported using more than 200,000 variants (including *APOE*) and a PGS area under the curve of 0.84 for Alzheimer’s disease (Escott-Price, Myers, Huentelman, & Hardy, 2017). Alzheimer’s disease has a strong genetic locus and the rest of the genome’s polygenic contribution has not been benchmarked across metrics of PGS construction.

Alzheimer’s disease PGS has not been assessed in population-based studies of dementia, other than by proxy in the UK Biobank cohort (Jansen et al.), and PGS construction metrics regarding the *APOE* region and additional SNPs have not been compared. The goals of this manuscript are three-fold. The first aim is to assess the utility of using polygenic scores in population-based analyses of dementia. The second aim is to evaluate the inclusion of the *APOE* region in these polygenic scores with and without a covariate modeling risk directly from the *APOE-ε4* allele. The third aim is to test SNP inclusion thresholds in PGS on dementia. We conduct this analysis using the Health and Retirement Study (HRS) in the European ancestries participants.

## Methods

### Health and Retirement Study

The Health and Retirement Study (HRS) is a nationally representative panel study featuring a biennial survey of adults over age 50 and their spouses in the United States (HRS, 2019). The HRS is sponsored by the National Institute on Aging (NIA U01AG009740) and is conducted by the University of Michigan. The HRS was established in 1992 as a means to provide a national resource for data on changing health and economic circumstances associated with ageing at both the individual and population levels. These changes are focused on four broads topics: income and wealth; health, cognition, and use of healthcare services; work and retirement; and family connections (Sonnega et al., 2014).

A random one-half of the sample was pre-selected to receive an enhanced face-to-face interview in 2006 which included physical performance tests, anthropometric measurements, blood and saliva samples, and a psychosocial self-administered questionnaire in addition to the HRS core interview. The remaining one-half sample was interviewed using the same enhanced face-to-face protocol in 2008. The new cohort in 2010 was also randomly assigned to receive an enhanced face-to-face interview in either 2010 or 2012. Those participants who were not interviewed or did not consent to saliva in 2006 were asked again in 2010.

Salivary DNA was collected using Oragene-250 saliva kits and protocol. DNA extracted from the saliva and was genotyped at the Center for Inherited Disease Research (CIDR) using the Illumina HumanOmni2.5 array (8v1 and 4v1). Genotyping Quality Control was performed by the Genetics Coordinating Center at the University of Washington, Seattle, WA. SNP annotation aligned to genome build 37/hg 19. Genetic principal components (PC) were calculated with HapMap controls (Patterson, Price, & Reich, 2006; Price et al., 2006). In addition to selecting independent SNPs with missing call rates < 5% and minor allele frequencies > 5%, the 2q21 (LCT), HLA, 8p23, and 17q21.31 regions were excluded from the initial pool (CIDR, 2013). The final European ancestries sample includes all self-reported non-Hispanic White persons that had PC loadings within ± one standard deviations of the mean for eigenvectors one and two in the PC analysis of all unrelated study subjects. Once the non-Hispanic White, European ancestries sample was identified, PCs were then re-calculated within this group to further account for population stratification. These “ancestry-specific principal components” were used in subsequent analyses. Imputation was performed using IMPUTE2 on HRS data phased using SHAPEIT2. Data were imputed to the 1000 Genomes Project (1000GP) cosmopolitan reference panel phase 3 version 5 (initial release on May 2013, haplotypes released Oct 2014) and are available on the database of genotypes and phenotypes (dbGaP, https://www.ncbi.nlm.nih.gov/gap/, phs000428.v2.p2).

### APOE-ε4

The apolipoprotein E (*APOE*) gene codes for a protein that binds and transports low-density lipids and is responsible, in part, for removing cholesterol from the bloodstream (Huang & Mahley, 2014; Mahley, 1988). Variations in this gene affect cholesterol metabolism and may lead to increases in the risk for stroke, heart disease, and may alter the odds of having Alzheimer’s disease. The *APOE* genotype is defined by two variants (rs7412 and rs429358) resulting in three common isoforms of *APOE*: *APOE-ε2, APOE-ε3*, and *APOE-ε4*. Of note, the genotyped rs7412 and rs429358 variants failed genotyping quality control in the HRS pipeline and are therefore not included as individual variants in any PGS using genotyped data alone. Using the imputed rs7412 (IMPUTE2 INFO score=0.988) and rs429358 (IMPUTE2 INFO score=0.99) variants, we categorized HRS participants as *ε2/ε2, ε2/ε3, ε2/ε4, ε3/ε3, ε3/ε4*, and *ε4/ε4*. We analyze two indicator variables for presence of one *ε4* allele (1=yes, 0=no) or two *ε4* allele (1=yes, 0=no).

## Polygenic Score for Alzheimer’s disease

We investigate using genome-wide raw genotyped variants and the two imputed *APOE* variants (rs7412 and rs429358) in the creation of our PGSs. We include SNPs in our PGS analysis at two AD-SNP association p-value thresholds (pT) from the Kunkle et al 2019 summary statistics: pT=0.01 and pT=1.0. PGSs at pT=0.01 include only those variants for which the association p-value in the IGAP meta-analysis on AD was less than 0.01. Summary statistics were obtained from National Institute on Aging Genetics of Alzheimer’s Disease Data Storage Site https://www.niagads.org/datasets/ng00075. Importantly, our study sample was not included in the Kunkle study of clinical Alzheimer’s disease. Thus, the weights are independent of our study sample. We do not include any linkage disequilibrium thresholding in our scores (i.e. no clumping or pruning algorithms). That is, we include any variants that pass quality control from the HRS and overlap with those variants from the AD GWAS summary statistics in our scores, unless otherwise noted.

For our scores with the *APOE* gene region removed, we removed all variants from the summary statistics on chromosome 19 (45,384,477 to 45,432,606, build 37/hg 19). This represents the start position of *TOMM40* (45,394,477) -10 kilobases and the stop position of *APOC1* (45,422,606) +10 kilobases. This region was removed in its entirety due to the dense linkage disequilibrium block in European ancestries overlapping these three genes (*TOMM40, APOE, APOC1*). We compare four PGSs: genotyped PGS including the *APOE* gene region, genotyped PGS with *APOE* region removed, each at two SNP p-values of significance pT=1 and pT< 0.01 (**Figure 1**).

**Figure 1.**
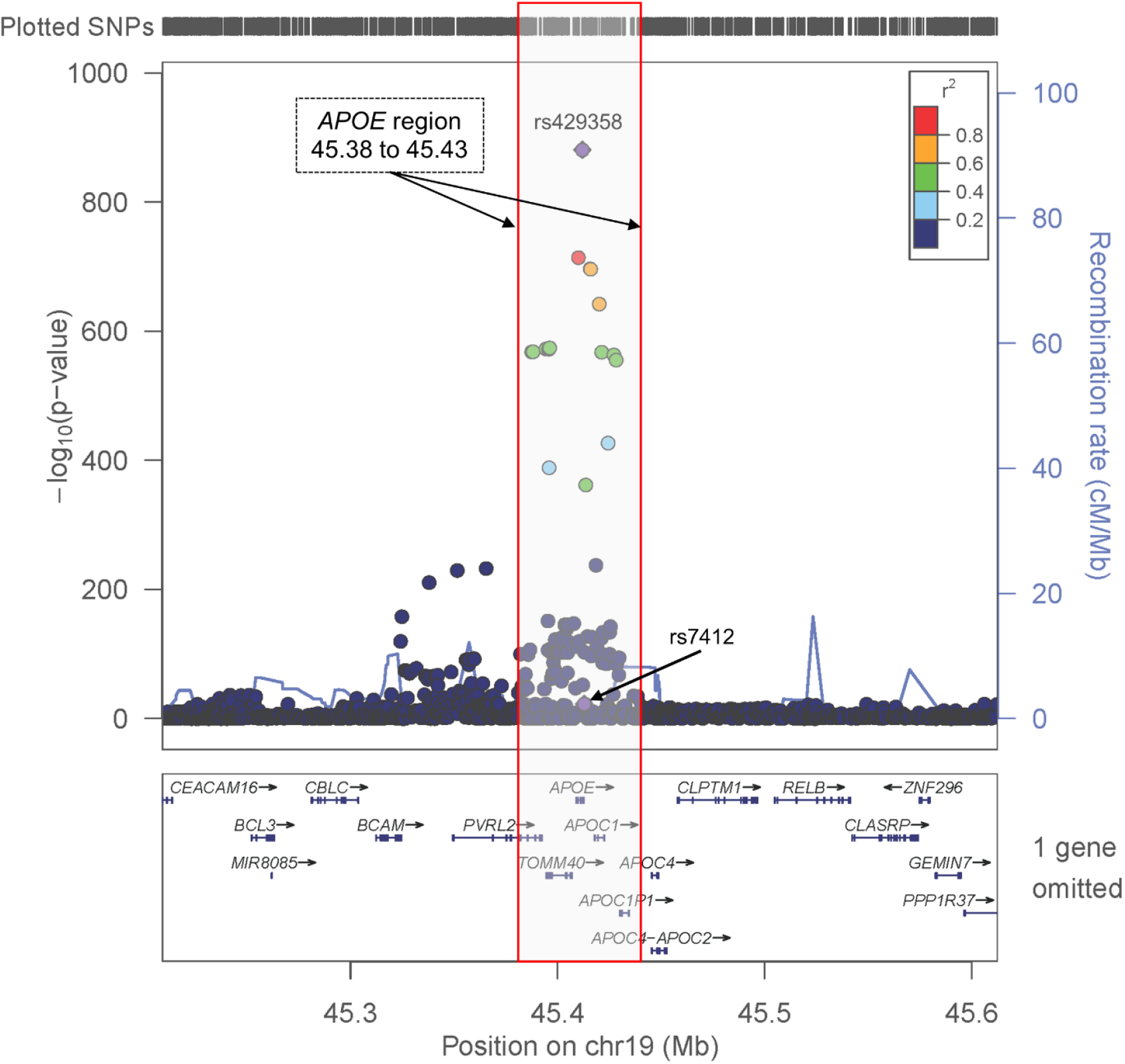
The *APOE/TOMM40* genomic locus on chromosome 19. The y-axis corresponds to - log_10_(p-values) of association with Alzheimer’s disease in Kunkle et al 2019. Single nucleotide polymorphisms within the bracketed genomic region were removed from consideration in polygenic score development for scores designed to exclude the *APOE* region. The variants that make the *APOE* isoforms are highlighted in purple diamond (rs7412, rs429358).

## Cognition Status

The HRS used a multidimensional measure of cognitive functioning, based on a telephone screen named the Telephone Interview for Cognitive Status (Brandt, Spencer, & Folstein, 1998). Domains assessed using this measure include: memory, mental status, abstract reasoning, fluid reasoning, vocabulary, dementia, and numeracy. In 2009, Langa, Kabeto, and Weir developed an approach for defining dementia and cognitively impaired non-dementia (CIND) in the HRS. This method has been clinically validated using equipercentile equating in the HRS against the Aging, Demographics, and Memory Study (ADAMS) – a sub-sample of the HRS who received a more extensive neurological battery and was evaluated by a team of dementia experts (Langa, Kabeto, & Weir, 2009; Langa et al., 2005). For self-respondents, the score consists of overall cognitive test performance while the proxy respondents’ scores are composed of proxy-rated memory, interviewer-perceived cognition, and impaired activities of daily living. The cut points for this method reflect the prevalence of dementia or cognitive impairment to the expected population prevalence from the ADAMS study. We used repeated measures of the classification of cognitive function data contributed for assessment years 2000

– 2014 from the HRS imputed cognition researcher contribution data set (Langa, Weir, Kabeto, & Sonnega, November 2018). For self-respondents, a score from 0 to 6 is categorized as dementia, 7 to 11 is categorized as cognitive impaired not dementia, and 12 to 27 is categorized as normal cognition. For proxy respondents starting in 2000, a score of 6 or higher out of 11 is classified as having dementia, a score of 3 to 5 indicates cognitive impaired not dementia, while 0 to 2 indicates normal cognition (Langa et al., 2009). In this analysis, we are only testing the odds of dementia versus normal cognition (1=dementia, 0=normal cognition).

## Covariates

Education attainment (years of school), birth cohort (AHEAD: Asset and Health Dynamics Among the Oldest Old (b. <1924); CODA: Children of the Depression (b. 1924-1930); HRS: Health and Retirement Study – original cohort (b. 1931-1941); WB: War babies (b. 1942-1947); EBB: early baby boomers (b. 1948-1953); MBB: mid-baby boomers (b. 1954-1959)), and sex (0=female, 1=male) are measured at the baseline HRS exam. Age (years) and a self-report of doctor diagnosed stroke (0=none, 1=stroke, possible stroke/TIA/mini-strokes, respondent disputes previous waves that indicate condition) are assessed at the same wave as the corresponding cognition visit.

## Statistical analysis

Due to the repeated measures in this analysis, we use generalized linear models and the GENMOD procedure in SAS 9.4. We specify repeated measures on the individual, a binomial distribution, and a logit link with an unstructured correlation structure. Fixed effects covariates included in every model included sex, years of education, and five ancestry-specific principal components. The time varying covariates chronological age, year, and stroke history at each interview wave are also included in every model. We use an α of 0.05 as a threshold for significance.

## Results

There are 9,872 individuals in the HRS non-Hispanic White, European ancestry analytic sample collected between 2006 and 2010. We removed observations with missing cognition (m=10,958), observations where the cognitive status was classified as CIND (m=6,905), and observations with a missing stroke status (m=16). This removed 55 additional individuals from the analysis. The final analysis included n=9,817 HRS respondents of predominantly European ancestries with m=51,225 cognitive observations (**Figure 2**).

**Figure 2.**
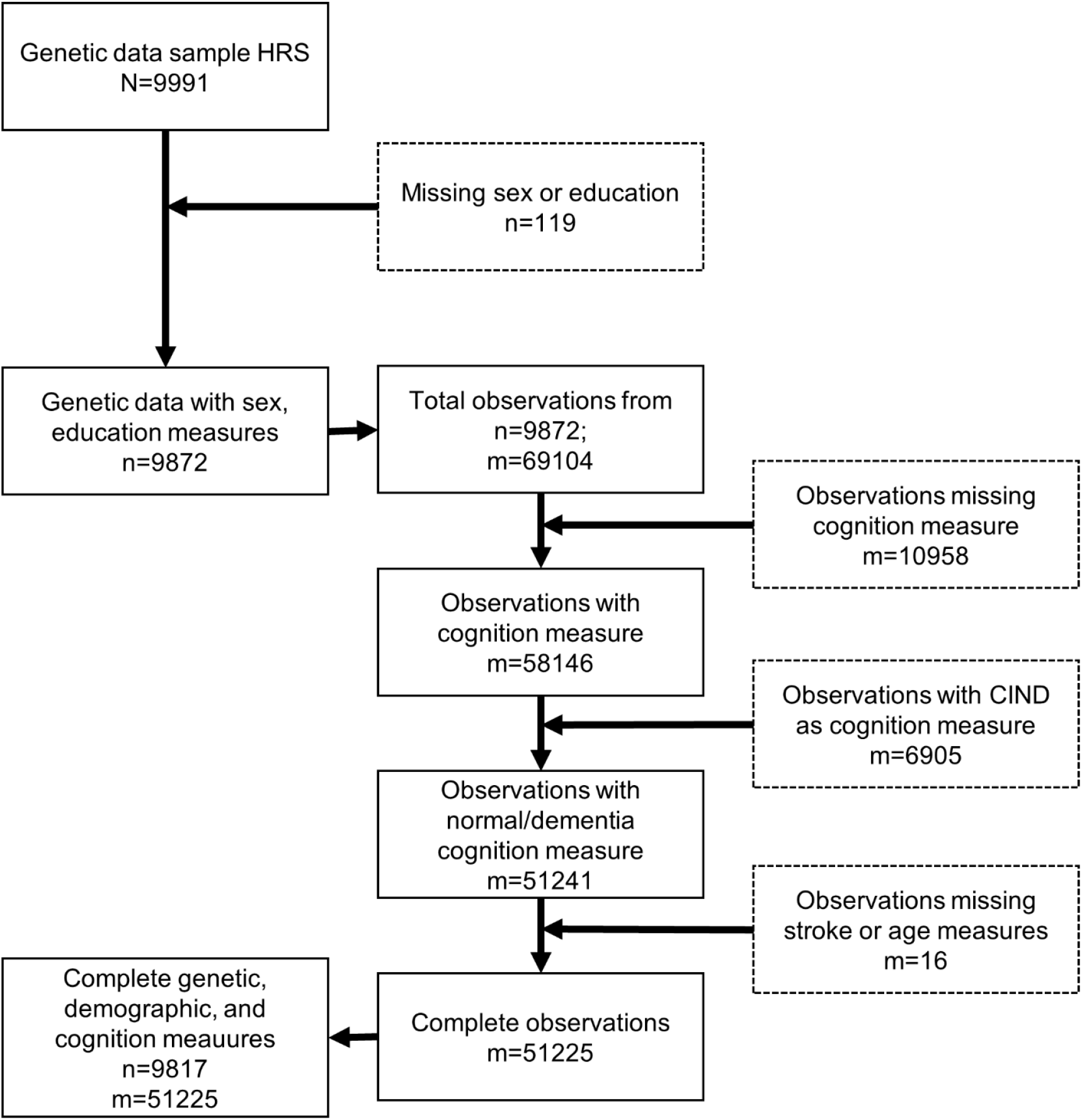
Sample inclusion flow chart for the Health and Retirement Study. HRS: Health and Retirement Study; n: number of individuals; m: num ber of observations; CIND: cognitively impaired, non-dementia

Our analytic sample was 57.8% female with an average age of 63.3 (SD=10.3) at their first visit during the observation period. On average, participants had 13.2 (2.5) years of education. The average number of cognitive assessments per participant was 5.2. A small portion of the sample had a history of stroke at their first visit during the observation period (n=417, 4.3%), with a higher proportion of males reporting a history of stroke at their first visit (n=201, 4.9%) than females (n=216, 3.8%). In unadjusted analyses, the AD PGS at either p-value threshold (pT) was not significantly different between males and females and broadly centered at zero with a standard deviation of one (pT=0.01, *P*=0.402; pT=1.0, *P*=0.71). There was no difference in the distribution of number of copies of *APOE-ε4* by sex, where the overall proportion with one copy of *APOE-ε4* was 24.3% (n=2382), and two copies of *APOE-ε4* was 2.1% (n=210). Across all observations, accounting for repeated measures within individuals, age, and dementia at first visit during the observation period were not significantly different by sex. The proportion of stroke; however, was significantly different by sex (*P*=0.0004) with a higher proportion in males than in females (**Table 1**). *APOE-ε4* status is associated (*Ps*<.05) with PGS (**Figure 3**).

**Table 1.**
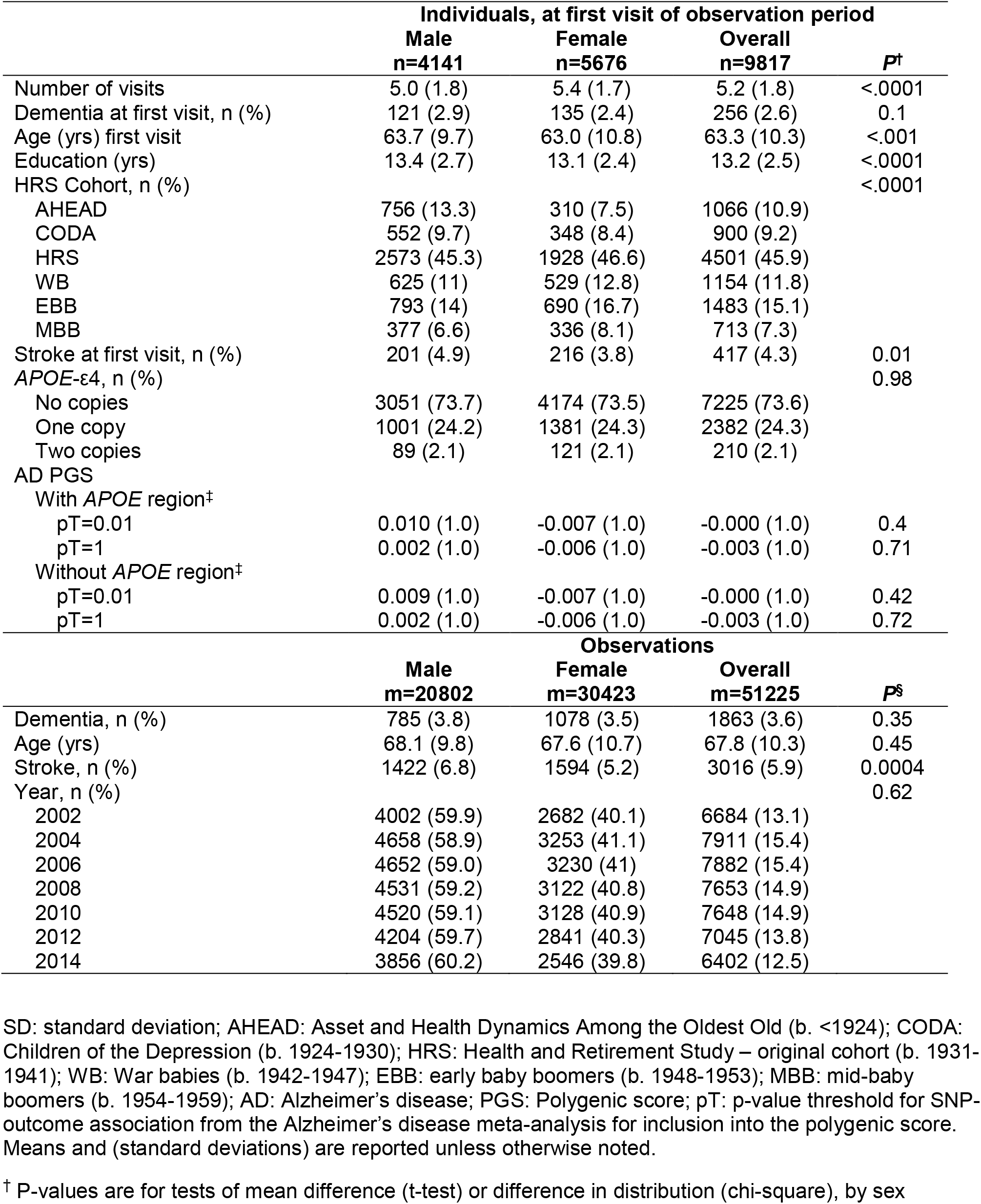
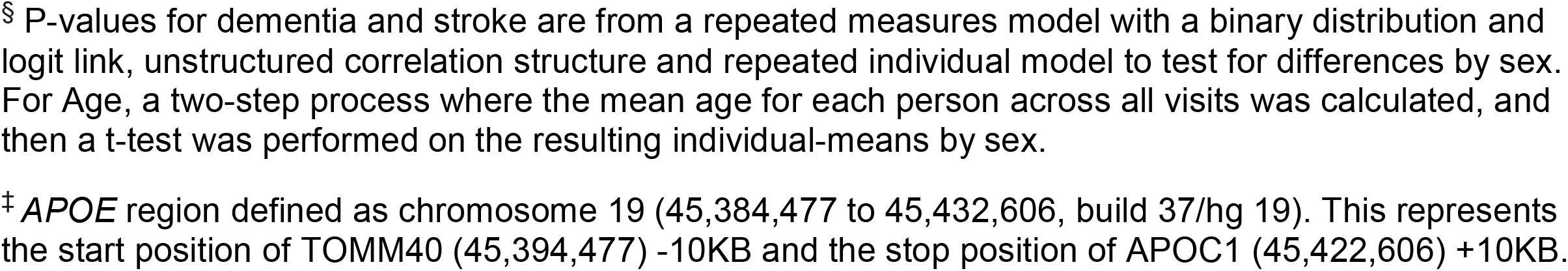
Individual- and observation-level descriptive statistics in the Health and Retirement Study

|  | Individuals, at first visit of observation period |  |  | *P*<sup>†</sup> |
| --- | --- | --- | --- | --- |
|  | Male<br>n=4141 | Female<br>n=5676 | Overall<br>n=9817 |  |
| Number of visits | 5.0 (1.8) | 5.4 (1.7) | 5.2 (1.8) | <.0001 |
| Dementia at first visit, n (%) | 121 (2.9) | 135 (2.4) | 256 (2.6) | 0.1 |
| Age (yrs) first visit | 63.7 (9.7) | 63.0 (10.8) | 63.3 (10.3) | <.001 |
| Education (yrs) | 13.4 (2.7) | 13.1 (2.4) | 13.2 (2.5) | <.0001 |
| HRS Cohort, n (%) |  |  |  | <.0001 |
| AHEAD | 756 (13.3) | 310 (7.5) | 1066 (10.9) |  |
| CODA | 552 (9.7) | 348 (8.4) | 900 (9.2) |  |
| HRS | 2573 (45.3) | 1928 (46.6) | 4501 (45.9) |  |
| WB | 625 (11) | 529 (12.8) | 1154 (11.8) |  |
| EBB | 793 (14) | 690 (16.7) | 1483 (15.1) |  |
| MBB | 377 (6.6) | 336 (8.1) | 713 (7.3) |  |
| Stroke at first visit, n (%) | 201 (4.9) | 216 (3.8) | 417 (4.3) | 0.01 |
| *APOE*-ε4, n (%) |  |  |  | 0.98 |
| No copies | 3051 (73.7) | 4174 (73.5) | 7225 (73.6) |  |
| One copy | 1001 (24.2) | 1381 (24.3) | 2382 (24.3) |  |
| Two copies | 89 (2.1) | 121 (2.1) | 210 (2.1) |  |
| AD PGS |  |  |  |  |
| With *APOE* region<sup>‡</sup> |  |  |  |  |
| pT=0.01 | 0.010 (1.0) | -0.007 (1.0) | -0.000 (1.0) | 0.4 |
| pT=1 | 0.002 (1.0) | -0.006 (1.0) | -0.003 (1.0) | 0.71 |
| Without *APOE* region<sup>‡</sup> |  |  |  |  |
| pT=0.01 | 0.009 (1.0) | -0.007 (1.0) | -0.000 (1.0) | 0.42 |
| pT=1 | 0.002 (1.0) | -0.006 (1.0) | -0.003 (1.0) | 0.72 |
|  | Observations |  |  | *P*<sup>§</sup> |
|  | Male<br>m=20802 | Female<br>m=30423 | Overall<br>m=51225 |  |
| Dementia, n (%) | 785 (3.8) | 1078 (3.5) | 1863 (3.6) | 0.35 |
| Age (yrs) | 68.1 (9.8) | 67.6 (10.7) | 67.8 (10.3) | 0.45 |
| Stroke, n (%) | 1422 (6.8) | 1594 (5.2) | 3016 (5.9) | 0.0004 |
| Year, n (%) |  |  |  | 0.62 |
| 2002 | 4002 (59.9) | 2682 (40.1) | 6684 (13.1) |  |
| 2004 | 4658 (58.9) | 3253 (41.1) | 7911 (15.4) |  |
| 2006 | 4652 (59.0) | 3230 (41) | 7882 (15.4) |  |
| 2008 | 4531 (59.2) | 3122 (40.8) | 7653 (14.9) |  |
| 2010 | 4520 (59.1) | 3128 (40.9) | 7648 (14.9) |  |
| 2012 | 4204 (59.7) | 2841 (40.3) | 7045 (13.8) |  |
| 2014 | 3856 (60.2) | 2546 (39.8) | 6402 (12.5) |  |

SD: standard deviation; AHEAD: Asset and Health Dynamics Among the Oldest Old (b. <1924); CODA: Children of the Depression (b. 1924-1930); HRS: Health and Retirement Study – original cohort (b. 1931-1941); WB: War babies (b. 1942-1947); EBB: early baby boomers (b. 1948-1953); MBB: mid-baby boomers (b. 1954-1959); AD: Alzheimer's disease; PGS: Polygenic score; pT: p-value threshold for SNP-outcome association from the Alzheimer's disease meta-analysis for inclusion into the polygenic score. Means and (standard deviations) are reported unless otherwise noted.
<sup>†</sup> P-values are for tests of mean difference (t-test) or difference in distribution (chi-square), by sex
§ P-values for dementia and stroke are from a repeated measures model with a binary distribution and logit link, unstructured correlation structure and repeated individual model to test for differences by sex. For Age, a two-step process where the mean age for each person across all visits was calculated, and then a t-test was performed on the resulting individual-means by sex.
‡ *APOE* region defined as chromosome 19 (45,384,477 to 45,432,606, build 37/hg 19). This represents the start position of TOMM40 (45,394,477) -10KB and the stop position of APOC1 (45,422,606) +10KB.

**Figure 3.**
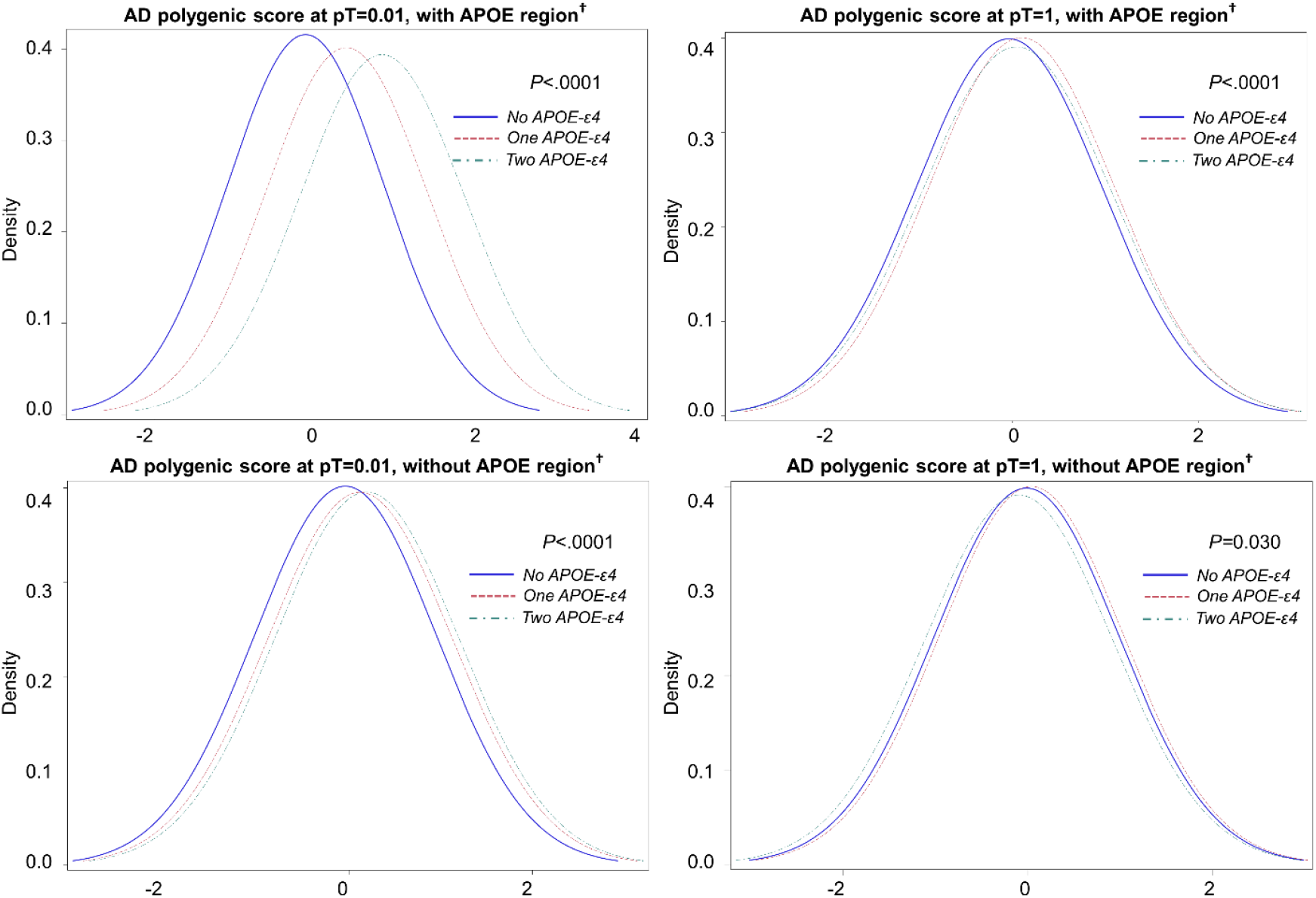
Distribution of Alzheimer’s disease polygenic score, by number of copies of *APOE-ε4* in the Health and Retirement Study, n=9871 AD: Alzheimer’s disease; pT: p-value threshold for SNP-outcome association from the Alzheimer’s disease meta-analysis for inclusion into the polygenic score. ^†^ *APOE* region defined as chromosome 19 (45,384,477 to 45,432,606, build 37/hg 19). This represents the start position of TOMM40 (45,394,477) -10KB and the stop position of APOC1 (45,422,606) +10KB.

In adjusted repeated measures regression models, increased age, later year of observation, history of stroke, and lower education were associated with increased odds of dementia in all models (all *P*<0.0001; **Table 2**). In the models using an AD PGS with the *APOE* region included, having one copy of *APOE-ε4* increased the odds of dementia relative to normal cognition by roughly 2 fold (AD PGS pT=0.01, OR=2.1 95%CI 1.8 to 2.5; AD PGS pT=1, OR=2.2 95%CI 1.9 to 2.6; **Table 2**), while two copies of *APOE-ε4* increased the odds of dementia by over 4 fold (AD PGS pT=0.01, OR=4.5 95%CI 2.9 to 6.9]; AD PGS pT=1, OR=4.9 95%CI 3.2 to 7.5; **Table 2**), holding all other variables constant. The AD PGS with the *APOE* region included was marginally associated with the odds of dementia relative to normal cognition only for the pT cutoff of 0.01 (OR=1.1 95%CI 1.0 to 1.2), while the AD PGS at pT=1 was not significantly associated with the odds of dementia. In the models that utilized the AD PGS with the *APOE* region removed, we observed similar estimates for one and two copies of *APOE-ε4* as before, and a similar 9% increase in the odds of dementia for every one standard deviation increase in the AD PGS at pT=0.01 (OR=1.1 95%CI 1.0 to 1.2). The AD PGS at pT=1 was not associated with the odds of dementia, relative to normal cognition in this sample.

**Table 2.** Odds ratios from repeated measures generalized linear models regression analyses of dementia in the Health and Retirement Study, n=9871, m=51225.

| Polygenic score with *APOE* region† |  |  |  |  |  |  |
| --- | --- | --- | --- | --- | --- | --- |
|  | pT=0.01 |  |  | pT=1.0 |  |  |
|  | OR (SE) | 95% CI | P | OR (SE) | 95% CI | P |
| Age (yrs) | 1.15 (1.01) | [1.14, 1.16] | <.0001 | 1.15 (1.01) | [1.14, 1.16] | <.0001 |
| Sex (male) | 1.13 (1.08) | [0.96, 1.32] | 0.132 | 1.12 (1.08) | [0.96, 1.32] | 0.149 |
| Educational (yrs) | 0.76 (1.02) | [0.74, 0.79] | <.0001 | 0.76 (1.02) | [0.74, 0.79] | <.0001 |
| Year | 1.12 (1.01) | [1.10, 1.14] | <.0001 | 1.12 (1.01) | [1.10, 1.14] | <.0001 |
| Stroke | 3.37 (1.10) | [2.82, 4.02] | <.0001 | 3.37 (1.10) | [2.82, 4.03] | <.0001 |
| No copies of *APOE*-ε4 | - | - | - | - | - | - |
| One copy of *APOE*-ε4 | 2.12 (1.09) | [1.78, 2.53] | <.0001 | 2.20 (1.09) | [1.85, 2.62] | <.0001 |
| Two copies of *APOE*-ε4 | 4.48 (1.25) | [2.90, 6.94] | <.0001 | 4.86 (1.25) | [3.16, 7.48] | <.0001 |
| AD polygenic score | 1.08 (1.04) | [1.00, 1.17] | 0.064 | 1.06 (1.05) | [0.97, 1.16] | 0.166 |
| Polygenic score without *APOE* region† |  |  |  |  |  |  |
|  | pT=0.01 |  |  | pT=1.0 |  |  |
|  | OR (SE) | 95% CI | P | OR (SE) | 95% CI | P |
| Age (yrs) | 1.15 (1.01) | [1.14, 1.16] | <.0001 | 1.15 (1.01) | [1.14, 1.16] | <.0001 |
| Sex (male) | 1.13 (1.08) | [0.97, 1.32] | 0.129 | 1.12 (1.08) | [0.96, 1.32] | 0.149 |
| Educational (yrs) | 0.76 (1.02) | [0.74, 0.79] | <.0001 | 0.76 (1.02) | [0.74, 0.79] | <.0001 |
| Year | 1.12 (1.01) | [1.10, 1.14] | <.0001 | 1.12 (1.01) | [1.10, 1.14] | <.0001 |
| Stroke | 3.37 (1.10) | [2.82, 4.02] | <.0001 | 3.37 (1.10) | [2.82, 4.02] | <.0001 |
| No copies of *APOE*-ε4 | - | - | - | - | - | - |
| One copy of *APOE*-ε4 | 2.18 (1.09) | [1.83, 2.59] | <.0001 | 2.21 (1.09) | [1.86, 2.63] | <.0001 |
| Two copies of *APOE*-ε4 | 4.72 (1.24) | [3.07, 7.25] | <.0001 | 4.92 (1.25) | [3.20, 7.56] | <.0001 |
| AD polygenic score | 1.09 (1.04) | [1.01, 1.18] | 0.028 | 1.07 (1.05) | [0.98, 1.16] | 0.144 |

OR: Odds ratio estimate for dementia relative to normal cognition; SE: standard error; CI: confidence interval; P: p-value for the OR estimate; *APOE*: Apolipoprotein E; AD: Alzheimer's disease; pT: p-value threshold for SNP-outcome association from the Alzheimer's disease meta-analysis for inclusion into the polygenic score.
All models were additionally adjusted for five ancestry-specific principal components. Generalized linear models accounted for repeated measures by individual and used a binomial distribution, logit link, and unstructured correlation structure.
† *APOE* region defined as chromosome 19 (45,384,477 to 45,432,606, build 37/hg 19). This represents the start position of TOMM40 (45,394,477) -10KB and the stop position of APOC1 (45,422,606) +10KB.

Because it has been demonstrated that there is mortality selection in the oldest individuals in the HRS genetic sample (Domingue et al., 2017), we removed the two oldest cohorts (AHEAD and CODA) as a sensitivity analysis (**Supplementary Table 1**). A total of 2,020 individuals and 8,957 observations were removed. The effect of any copy of *APOE-ε4* remained highly associated with the odds of dementia compared to normal cognition. Though the effect size for the AD PGSs at each pT and whether or not the *APOE* region were not significantly different than those in **Table 2**, the p-values associated with these effects were non-significant for all AD PGS. The slightly attenuated effects are not surprising as the younger cohorts are just now entering ages at which dementia becomes more prevalent.

## Discussion

In a large, population-based cohort of older, European ancestry Americans, cumulative genetic risk summarized as a PGS is informative of longitudinal dementia risk. We observed that the *APOE* region requires handling with care in the development of PGS. Specifically, including the *APOE* region as weighted SNPs in a PGS was insufficient to account for the large risk attributed to the *APOE* region. We recommend removing the region in linkage disequilibrium around the *APOE* locus from the PGS and treating the *APOE* locus as an independent covariate. In addition, we observed greater performance from PGS developed with a more stringent threshold p-value for SNP inclusion, with greater noise from a PGS informed by the full genome in association with this dementia phenotype. Optimized measures of the polygenic nature of dementia allow for more powerful interrogations of genetic and environmental risk for dementia.

We observed the *APOE-ε4* allele was longitudinally associated with higher risk of dementia, in a dose dependent manner. This observation is consistent with extensive prior research (Logue et al., 2019; Tanzi & Bertram, 2001). The *APOE-ε4* allele is neither necessary, nor sufficient to cause dementia, but the magnitude of increased risk attributed to each copy of the allele is relatively high. The *APOE-ε4* allele is in linkage disequilibrium with a 100 kilobase region involving the *APOE, APOEC*, and *TOMM40* genes. Thus, an *APOE* independent PGS would need to remove the SNPs from the entire *APOE* region. In excess of the association between the *APOE* region and dementia, we observed a small, but significant association between AD PGS and dementia. These findings are similar to those observed in clinical populations investigating *APOE* independent PGS risk of Alzheimer’s disease specifically (Cruchaga et al., 2018; Desikan et al., 2017; Escott-Price et al., 2017; Lupton et al., 2016; Tan et al., 2018). When building PGS, it is important to have independent study samples between the discovery GWAS and the application PGS. Notably, our study sample was part of the Kunkle GWAS that generated the weights for the PGS. Our findings show the *APOE* independent AD PGS can be successfully implemented in population-based research of a broad dementia phenotype.

Dementia is a disorder with a strong genetic locus of effect (*APOE*) and substantially weaker effects are scattered throughout the genome. Including the *APOE* region in PGS without specific measurement of *APOE-ε4* is insufficient, and overestimates the polygenic nature of dementia. Similarly, in Amyotrophic Lateral Sclerosis (ALS) there is a strong main effect locus (*C9orf72*), and a significant, albeit modest, proportion of the phenotypic variance could be explained by polygenic risk score, over and above the *C9orf72* region (van Rheenen et al., 2016). In contrast, other chronic disease traits, such as obesity, lack a dominant genetic locus and polygenic score development is successful across the entire genome at a higher p-value threshold (Ware et al., 2017). Together, these results suggest that in traits with a strong genetic locus, polygenic scores should exclude the primary regions and seek to aggregate the remaining genetic risk as a separate predictor.

Our research has several limitations that should be acknowledged. The first is that our study relied on imputed *APOE* variant calls. The *APOE* region is notoriously difficult and labor intensive to measure genotypes (Radmanesh et al., 2014; Zhong et al., 2016). Indeed, the two primary *APOE* SNPs of interest failed quality control metrics on the genotyping array in the HRS. We used the correlation structure of the genome from the 1000GP reference to impute these SNPs with ∼99% confidence. Second, our study may be subject to mortality selection; however, dementia is primarily a disease of older age and requires survival long enough to manifest symptoms. Mortality selection related to the *APOE* genotype would only serve to make our observations more conservative. Third, our PGS was developed using weights from a GWAS of primarily European ancestry participants, thereby limiting generalizability to other acnestries. Last, our population-based study assessed a broad phenotype of dementia. There are many types of dementia including Alzheimer’s, vascular, and frontotemporal lobe, which have varying genetic architectures, to which we applied a PGS specific for Alzheimer’s. As future GWAS become available for dementia subtypes in a clinical population, investigators may be able to classify the utility of PGS in dementia subtypes.

Dementia has considerable risk attributed to genetic factors. The *APOE* region is the strongest locus associated with disease and many additional sites confer small risk effects. Incorporating genetic risk from many sites in a polygenic risk score is a useful metric for risk prediction and etiologic testing in epidemiologic research of complex traits (Maher, 2015). Our findings demonstrate the *APOE* region should be removed prior to polygenic risk score development and treated as an independent factor in dementia analyses. More work is needed to assess polygenic scores for Alzheimer’s disease for clinical utility and prediction and in diverse ancestries.

## Data Availability

All data from these analyses are available through the Health and Retirement Study (https://hrs.isr.umich.edu/) or the database of genotypes and phenotypes (dbGaP https://www.ncbi.nlm.nih.gov/gap/, phs000428.v2.p2).

https://www.niagads.org/datasets/ng00075

https://www.ncbi.nlm.nih.gov/gap/

https://hrs.isr.umich.edu/

## Acknowledgments

The authors would like to thank the participants of the Health and Retirement Study.

---

**Supplementary Table 1.**
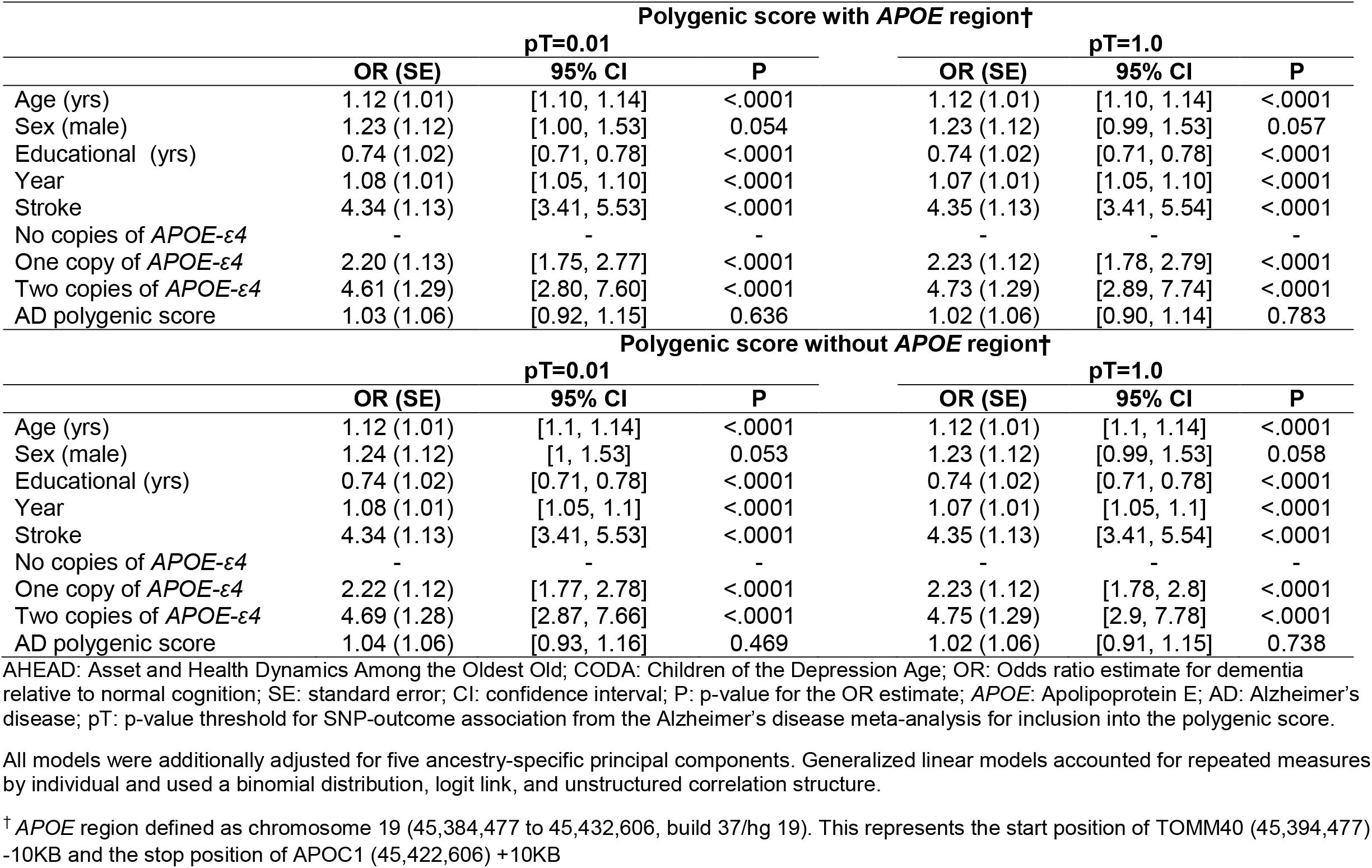
Odds ratios from repeated measures generalized linear models regression analyses of dementia removing AHEAD and CODA in the Health and Retirement Study, n=7851, m=42268.

| Polygenic score with *APOE* region† |  |  |  |  |  |  |
| --- | --- | --- | --- | --- | --- | --- |
|  | pT=0.01 |  |  | pT=1.0 |  |  |
|  | OR (SE) | 95% CI | P | OR (SE) | 95% CI | P |
| Age (yrs) | 1.12 (1.01) | [1.10, 1.14] | <.0001 | 1.12 (1.01) | [1.10, 1.14] | <.0001 |
| Sex (male) | 1.23 (1.12) | [1.00, 1.53] | 0.054 | 1.23 (1.12) | [0.99, 1.53] | 0.057 |
| Educational (yrs) | 0.74 (1.02) | [0.71, 0.78] | <.0001 | 0.74 (1.02) | [0.71, 0.78] | <.0001 |
| Year | 1.08 (1.01) | [1.05, 1.10] | <.0001 | 1.07 (1.01) | [1.05, 1.10] | <.0001 |
| Stroke | 4.34 (1.13) | [3.41, 5.53] | <.0001 | 4.35 (1.13) | [3.41, 5.54] | <.0001 |
| No copies of *APOE*-ε4 | - | - | - | - | - | - |
| One copy of *APOE*-ε4 | 2.20 (1.13) | [1.75, 2.77] | <.0001 | 2.23 (1.12) | [1.78, 2.79] | <.0001 |
| Two copies of *APOE*-ε4 | 4.61 (1.29) | [2.80, 7.60] | <.0001 | 4.73 (1.29) | [2.89, 7.74] | <.0001 |
| AD polygenic score | 1.03 (1.06) | [0.92, 1.15] | 0.636 | 1.02 (1.06) | [0.90, 1.14] | 0.783 |
| Polygenic score without *APOE* region† |  |  |  |  |  |  |
|  | pT=0.01 |  |  | pT=1.0 |  |  |
|  | OR (SE) | 95% CI | P | OR (SE) | 95% CI | P |
| Age (yrs) | 1.12 (1.01) | [1.1, 1.14] | <.0001 | 1.12 (1.01) | [1.1, 1.14] | <.0001 |
| Sex (male) | 1.24 (1.12) | [1, 1.53] | 0.053 | 1.23 (1.12) | [0.99, 1.53] | 0.058 |
| Educational (yrs) | 0.74 (1.02) | [0.71, 0.78] | <.0001 | 0.74 (1.02) | [0.71, 0.78] | <.0001 |
| Year | 1.08 (1.01) | [1.05, 1.1] | <.0001 | 1.07 (1.01) | [1.05, 1.1] | <.0001 |
| Stroke | 4.34 (1.13) | [3.41, 5.53] | <.0001 | 4.35 (1.13) | [3.41, 5.54] | <.0001 |
| No copies of *APOE*-ε4 | - | - | - | - | - | - |
| One copy of *APOE*-ε4 | 2.22 (1.12) | [1.77, 2.78] | <.0001 | 2.23 (1.12) | [1.78, 2.8] | <.0001 |
| Two copies of *APOE*-ε4 | 4.69 (1.28) | [2.87, 7.66] | <.0001 | 4.75 (1.29) | [2.9, 7.78] | <.0001 |
| AD polygenic score | 1.04 (1.06) | [0.93, 1.16] | 0.469 | 1.02 (1.06) | [0.91, 1.15] | 0.738 |

AHEAD: Asset and Health Dynamics Among the Oldest Old; CODA: Children of the Depression Age; OR: Odds ratio estimate for dementia relative to normal cognition; SE: standard error; CI: confidence interval; P: p-value for the OR estimate; *APOE*: Apolipoprotein E; AD: Alzheimer's disease; pT: p-value threshold for SNP-outcome association from the Alzheimer's disease meta-analysis for inclusion into the polygenic score.
All models were additionally adjusted for five ancestry-specific principal components. Generalized linear models accounted for repeated measures by individual and used a binomial distribution, logit link, and unstructured correlation structure.
† *APOE* region defined as chromosome 19 (45,384,477 to 45,432,606, build 37/hg 19). This represents the start position of TOMM40 (45,394,477) -10KB and the stop position of APOC1 (45,422,606) +10KB

## Notes

**Funding** This work was supported by grants from the National Institutes of Health (NIA R01 AG055406, NIA RF1 AG055654, NIMHD R01 MD011716).

### Competing Interest Statement

The authors have declared no competing interest.

### Funding Statement

This work was supported by grants from the National Institutes of Health (NIA R01 AG055406, NIA RF1 AG055654, NIMHD R01 MD011716).

## References

Brandt, J., Spencer, M., & Folstein, M. (1998). The telephone interview for cognitive status. Neuropsychiatry Neuropsychol Behav Neurol(1), 111–117.

CIDR, C. f. I. D. R. (2013). Quality Control Report for Genotypic Data - Health and Retirement Study Phase 1–3. Retrieved from

Cruchaga, C., Del-Aguila, J. L., Saef, B., Black, K., Fernandez, M. V., Budde, J., … Harari, O. (2018). Polygenic risk score of sporadic late-onset Alzheimer’s disease reveals a shared architecture with the familial and early-onset forms. Alzheimers Dement, 14(2), 205–214. doi:10.1016/j.jalz.2017.08.013

Desikan, R. S., Fan, C. C., Wang, Y., Schork, A. J., Cabral, H. J., Cupples, L. A., … Dale, A. M. (2017). Genetic assessment of age-associated Alzheimer disease risk: Development and validation of a polygenic hazard score. PLoS Med, 14(3), e1002258. doi:10.1371/journal.pmed.1002258

Domingue, B. W., Belsky, D. W., Harrati, A., Conley, D., Weir, D. R., & Boardman, J. D. (2017). Mortality selection in a genetic sample and implications for association studies. Int J Epidemiol, 46(4), 1285–1294. doi:10.1093/ije/dyx041

Escott-Price, V., Myers, A. J., Huentelman, M., & Hardy, J. (2017). Polygenic risk score analysis of pathologically confirmed Alzheimer disease. Ann Neurol, 82(2), 311–314. doi:10.1002/ana.24999

Gatz, M., Pedersen, N. L., Berg, S., Johansson, B., Johansson, K., Mortimer, J. A., … Ahlbom, A. (1997). Heritability for Alzheimer’s disease: the study of dementia in Swedish twins. J Gerontol A Biol Sci Med Sci, 52(2), M117–125. doi:10.1093/gerona/52a.2.m117

Gatz, M., Reynolds, C. A., Fratiglioni, L., Johansson, B., Mortimer, J. A., Berg, S., … Pedersen, N. L. (2006). Role of genes and environments for explaining Alzheimer disease. Arch Gen Psychiatry, 63(2), 168–174. doi:10.1001/archpsyc.63.2.168

Harold, D., Abraham, R., Hollingworth, P., Sims, R., Gerrish, A., Hamshere, M. L., … Williams, J. (2009). Genome-wide association study identifies variants at CLU and PICALM associated with Alzheimer’s disease. Nat Genet, 41(10), 1088–1093. doi:10.1038/ng.440

HRS. (2019). Health and Retirement Study, 2004 core public use dataset.

Huang, Y., & Mahley, R. W. (2014). Apolipoprotein E: structure and function in lipid metabolism, neurobiology, and Alzheimer’s diseases. Neurobiol Dis, 72 Pt A, 3–12. doi:10.1016/j.nbd.2014.08.025

International Schizophrenia, C., Purcell, S. M., Wray, N. R., Stone, J. L., Visscher, P. M., O’Donovan, M. C., … Sklar, P. (2009). Common polygenic variation contributes to risk of schizophrenia and bipolar disorder. Nature, 460(7256), 748–752. doi:10.1038/nature08185

Jansen, I. E., Savage, J. E., Watanabe, K., Bryois, J., Williams, D. M., Steinberg, S., … Posthuma, D. (2019). Genome-wide meta-analysis identifies new loci and functional pathways influencing Alzheimer’s disease risk. Nat Genet, 51(3), 404–413. doi:10.1038/s41588-018-0311-9

Kunkle, B. W., Grenier-Boley, B., Sims, R., Bis, J. C., Damotte, V., Naj, A. C., … Environmental Risk for Alzheimer’s Disease, C. (2019). Genetic meta-analysis of diagnosed Alzheimer’s disease identifies new risk loci and implicates Abeta, tau, immunity and lipid processing. Nat Genet, 51(3), 414–430. doi:10.1038/s41588-019-0358-2

Lambert, J. C., Ibrahim-Verbaas, C. A., Harold, D., Naj, A. C., Sims, R., Bellenguez, C., … Amouyel, P. (2013). Meta-analysis of 74,046 individuals identifies 11 new susceptibility loci for Alzheimer’s disease. Nat Genet, 45(12), 1452–1458. doi:10.1038/ng.2802

Langa, K. M., Kabeto, M., & Weir, D. (2009). Report on race and cognitive impairment using HRS in 2010. Alzheimer’s disease facts and figures.

Langa, K. M., Plassman, B. L., Wallace, R. B., Herzog, A. R., Heeringa, S. G., Ofstedal, M. B., … Willis, R. J. (2005). The Aging, Demographics, and Memory Study: study design and methods. Neuroepidemiology, 25(4), 181–191. doi:10.1159/000087448

Langa, K. M., Weir, D., Kabeto, M., & Sonnega, A. (November 2018). Researcher Contribution: Langa-Weir Classification of Cognitive Function. Survey Research Center.

Liu, C. C., Liu, C. C., Kanekiyo, T., Xu, H., & Bu, G. (2013). Apolipoprotein E and Alzheimer disease: risk, mechanisms and therapy. Nat Rev Neurol, 9(2), 106–118. doi:10.1038/nrneurol.2012.263

Logue, M. W., Panizzon, M. S., Elman, J. A., Gillespie, N. A., Hatton, S. N., Gustavson, D. E., … Lyons, M. J. (2019). Use of an Alzheimer’s disease polygenic risk score to identify mild cognitive impairment in adults in their 50s. Molecular psychiatry, 24(3), 421.

Lupton, M. K., Strike, L., Hansell, N. K., Wen, W., Mather, K. A., Armstrong, N. J., … Wright, M. J. (2016). The effect of increased genetic risk for Alzheimer’s disease on hippocampal and amygdala volume. Neurobiol Aging, 40, 68–77. doi:10.1016/j.neurobiolaging.2015.12.023

Maher, B. S. (2015). Polygenic Scores in Epidemiology: Risk Prediction, Etiology, and Clinical Utility. Curr Epidemiol Rep, 2(4), 239–244. doi:10.1007/s40471-015-0055-3

Mahley, R. W. (1988). Apolipoprotein E: cholesterol transport protein with expanding role in cell biology. Science, 240(4852), 622–630. doi:10.1126/science.3283935

Martin, A. R., Daly, M. J., Robinson, E. B., Hyman, S. E., & Neale, B. M. (2019). Predicting Polygenic Risk of Psychiatric Disorders. Biol Psychiatry, 86(2), 97–109. doi:10.1016/j.biopsych.2018.12.015

Naj, A. C., Jun, G., Beecham, G. W., Wang, L. S., Vardarajan, B. N., Buros, J., … Schellenberg, G. D. (2011). Common variants at MS4A4/MS4A6E, CD2AP, CD33 and EPHA1 are associated with late-onset Alzheimer’s disease. Nat Genet, 43(5), 436–441. doi:10.1038/ng.801

Patterson, N., Price, A. L., & Reich, D. (2006). Population structure and eigenanalysis. PLoS Genet, 2(12), e190. doi:10.1371/journal.pgen.0020190

Price, A. L., Patterson, N. J., Plenge, R. M., Weinblatt, M. E., Shadick, N. A., & Reich, D. (2006). Principal components analysis corrects for stratification in genome-wide association studies. Nature genetics, 38%, 904. doi:10.1038/ng1847

https://www.nature.com/articles/ng1847#supplementary-information

Radmanesh, F., Devan, W. J., Anderson, C. D., Rosand, J., Falcone, G. J., & Alzheimer’s Disease Neuroimaging, I. (2014). Accuracy of imputation to infer unobserved APOE epsilon alleles in genome-wide genotyping data. Eur J Hum Genet, 22(10), 1239–1242. doi:10.1038/ejhg.2013.308

Seshadri, S., Fitzpatrick, A. L., Ikram, M. A., DeStefano, A. L., Gudnason, V., Boada, M., … Consortium, E. (2010). Genome-wide analysis of genetic loci associated with Alzheimer disease. JAMA, 303(18), 1832–1840. doi:10.1001/jama.2010.574

Sonnega, A., Faul, J., Ofstedal, M., Langa, K., Phillips, J., & Weir, D. (2014). Cohort Profile: the Health and Retirement Study (HRS). Int J Epidemiol, 43(2), 576–585.

Tan, C. H., Fan, C. C., Mormino, E. C., Sugrue, L. P., Broce, I. J., Hess, C. P., … Alzheimer’s Disease Neuroimaging, I. (2018). Polygenic hazard score: an enrichment marker for Alzheimer’s associated amyloid and tau deposition. Acta Neuropathol, 135(1), 85–93. doi:10.1007/s00401-017-1789-4

Tanzi, R. E., & Bertram, L. (2001). New Frontiers in Alzheimer’s Disease Genetics. Neuron, 32(2), 181–184. doi:https://doi.org/10.1016/S0896-6273(01)00476-7

Tosto, G., Bird, T. D., Tsuang, D., Bennett, D. A., Boeve, B. F., Cruchaga, C., … Mayeux, R. (2017). Polygenic risk scores in familial Alzheimer disease. Neurology, 88(12), 1180–1186. doi:10.1212/WNL.0000000000003734

Van Cauwenberghe, C., Van Broeckhoven, C., & Sleegers, K. (2016). The genetic landscape of Alzheimer disease: clinical implications and perspectives. Genet Med, 18(5), 421–430. doi:10.1038/gim.2015.117

van Rheenen, W., Shatunov, A., Dekker, A. M., McLaughlin, R. L., Diekstra, F. P., Pulit, S. L., … Veldink, J. H. (2016). Genome-wide association analyses identify new risk variants and the genetic architecture of amyotrophic lateral sclerosis. Nat Genet, 48(9), 1043–1048. doi:10.1038/ng.3622

Ward, A., Crean, S., Mercaldi, C. J., Collins, J. M., Boyd, D., Cook, M. N., & Arrighi, H. M. (2012). Prevalence of apolipoprotein E4 genotype and homozygotes (APOE e4/4) among patients diagnosed with Alzheimer’s disease: a systematic review and meta-analysis. Neuroepidemiology, 38(1), 1–17. doi:10.1159/000334607

Ware, E. B., Schmitz, L. L., Faul, J. D., Gard, A., Mitchell, C., Smith, J. A., … Kardia, S. L. R. (2017). Heterogeneity in polygenic scores for common human traits. bioRxiv. doi:10.1101/106062

Zhong, L., Xie, Y. Z., Cao, T. T., Wang, Z., Wang, T., Li, X., … Chen, X. F. (2016). A rapid and cost-effective method for genotyping apolipoprotein E gene polymorphism. Mol Neurodegener, 11%, 2. doi:10.1186/s13024-016-0069-4

